# Dietary and other risk factors for cardiovascular disease analysed with Global Burden of Disease worldwide cohorts: lipid hypothesis versus fat-soluble vitamin hypothesis

**DOI:** 10.1101/2021.04.17.21255675

**Authors:** David K Cundiff, Chunyi Wu

**Affiliations:** Long Beach, California, USA; Area Specialist Lead in Epidemiology and Statistics, Michigan Medicine, Ann Arbor, Michigan, USA; Volunteer collaborators with the Institute of Health Metrics and Evaluation, Seattle, Washington, USA

## Abstract

**Background:** Debate about whether high intake of dietary saturated fatty acids (SFA) causes coronary heart disease (the lipid hypothesis) is ongoing.

**Methods:** Using worldwide Global Burden of Disease (GBD) data on cardiovascular disease deaths/100k/year, ages 15-69 years old in male and female cohorts (CVD) and dietary and other CVD risk factors, we formatted and population weighted data from 195 countries. The formatted rows of data (n=7846 cohorts) each represented about 1 million people, totaling about 7.8 billion people in 2020. We correlated CVD with dietary and other risk factors worldwide and in appropriate subsets. Outcome measures included CVD versus dietary and other risk factor correlations worldwide and in subsets.

**Findings:** After empirical data exploration, we defined a “fat-soluble vitamin variable” Kcal/day (FSVV) as, “FSVV=processed meat + red meat + fish + milk + poultry + eggs + (SFA + polyunsaturated fatty acids (PUFA) + trans fatty acids (TFA)) * 0.46 (all in Kcal/day).” Low density lipoprotein cholesterol mmol/L correlated strongly positively with FSVV worldwide (r=0.780, 95% CI 0.771 to 0.788, p<0001, n=7846 cohorts), so we considered FSVV our marker variable to test both the lipid hypothesis and our fat-soluble vitamin hypothesis. LDL-C correlated negatively with CVD worldwide (r= -0.279, 95% CI -0.299 to -0.258, *p*<0.0001 as did FSVV versus CVD (r= -0.329, 95% CI -0.349 to -0.309, *p*<0.0001). However, FSVV correlated positively with CVD in the highest FSVV cohorts (mean male/female FSVV ≥567.27 Kcal/day: r=0.523, 95% CI 0.476 to 0.567, p<0001, n=974 cohorts).

**Interpretation:** Since FSVV correlated positively with CVD only in high FSVV cohorts, the data supported the lipid hypothesis only in GBD cohorts with high FSVV intake. Since both FSVV and LDL-C correlated negatively with CVD worldwide, only the fat-soluble vitamin hypothesis was supported worldwide. This GBD cohort data analysis methodology could be used to help develop food policy and education strategies for improving public health.

**Funding:** none

**Research in context:** *Evidence before this study:* Relating to dietary and other risk factors for early death from cardiovascular diseases worldwide, the EAT-Lancet Commission proposed the “Planetary Health Diet” to reduce non-communicable diseases and mitigate climate change. The Planetary Health Diet has been controversial and did not achieve the goal of leading a worldwide “Great Food Transformation” towards a more plant-based diet. The comparative risk assessment (CRA) systematic literature review-based methodology used in drafting the Planetary Health Diet has no significant competitors as published methodologies used to parse health effects of worldwide dietary and other risk factors. No consensus exists about an optimal range of diets for optimum human health, including cardiovascular disease prevention/treatment.

*Added value of this study□:* Cardiovascular disease deaths/100k/year in 15-69-year-old males and females (CVD) worldwide correlated with dietary risk factors had two patterns based on the amounts of kilocalories/day (Kcal/day) of animal foods and added fats. We differentiated the two patterns by defining a “fat-soluble vitamin variable” Kcal/day (FSVV=processed meat + red meat + fish + milk + poultry + eggs + (added saturated fatty acids (SFA)+polyunsaturated fatty acids (PUFA)+trans fatty acids (TFA)) * 0.46 (all in Kcal/day). Worldwide, all nine risk factors comprising FSVV correlated negatively with CVD, but in high FSVV cohorts (FSVV≥567.27 Kcal/day), all nine risk factors in the FSVV correlated positively with CVD.

*Implications of all the available evidence:* Animal foods and added fats afforded significant protection from CVD worldwide. The data suggested that the fat-soluble vitamins in animal foods and added fat facilitating gut absorption may have been partially or entirely responsible for the worldwide protection from CVD. However, in high FSVV intake cohorts, SFA in food and in extracted added fats facilitating atherosclerosis may have partially or entirely accounted for the positive correlation of FSVV and CVD. The findings suggest that a yet not precisely defined moderate amount of animal food and added fat intake contributed to lower cardiovascular risk relative to very high or very low intakes.

## Introduction

The scientific validity of the Dietary Guidelines for Americans for 2015-2020, including guidelines based on the lipid hypothesis, was challenged by Journalist Nina Teicholz in the *BMJ*.^1^ The Center for Science in the Public Interest called for the *BMJ* to retract the article. Peer reviewers the *BMJ* selected to adjudicate the far-reaching dispute concluded that, “Teicholz’s criticisms of the methods used by Dietary Guidelines for Americans Committee are within the realm of scientific debate.”^2^ Currently, no methodology for relating cardiovascular disease events to food intake has been generally accepted as rigorous, replicable, and scientifically valid.

This paper will use population weighted, formatted, worldwide global burden of disease (GBD) data from the Institute of Health Metrics and Evaluation (IHME) to assess the impact of diet, metabolic risk factors, tobacco use, air pollution, and other risk factors on cardiovascular disease deaths/100k/year in male and female cohorts 15-69-years-old (CVD).

We were open to the possible support of the data for the lipid hypothesis but also for our newly defined, “fat-soluble vitamin hypothesis,” which an initial exploration of the GBD data prompted. We defined it as “insufficient dietary intake of fat-soluble vitamin containing animal foods and added fat for gut absorption increases cardiovascular disease risk.” We will elaborate in the results and the discussion.

## Methods

As volunteer collaborators with the IHME, we acquired and utilised raw GBD worldwide ecological data (GBD: 195 countries and 365 subnational locations, n=1120 male and female cohorts). Data consisted of the rate of cardiovascular disease early deaths, metabolic risk factors, dietary risk factors and covariates, and other risk factors of male and female cohorts 15-49 years old and 50-69 years old from each year 1990-2017. GBD worldwide citations of over 12,000 surveys constituting ecological data inputs for this analysis are available online from IHME.^3^ The main characteristics of IHME GBD data sources, the protocol for the GBD study, and all risk factor values have been published by IHME GBD data researchers and discussed elsewhere.^4–8^ These include detailed descriptions of categories of input data, potentially important biases, and the methodologies of analysis. We did not clean or pre-process any of the GBD data. GBD cohort risk factor and health outcome data from the IHME had no missing records.

Food risk factors came from surveys that IHME utilised as gram/day (g/d) consumed on average. GBD dietary covariate data originally came from Food and Agriculture Organization surveys of animal and plant food commodities available per capita in countries worldwide (potatoes, corn, rice, sweet potatoes, poultry, and eggs)—as opposed to consumed on average.^9^

Supplementary Table 1 lists the relevant GBD dietary and other risk factors, covariates, and other available variables with definitions of those risk factor exposures.^7^ Once IHME staff time constraints due to COVID-19 data collection and modeling are over, the updated GBD 2020 data with all the variables as the GBD 2017 data that we used for this analysis may be obtained by volunteer researchers collaborating with IHME.^10^

### Study design and population

For cardiovascular disease early deaths and all risk factors, we averaged the values for ages 15-49 years old together with 50-69 years old for each male and female cohort for each year. Finally, for each male and female cohort, data from all 28 years (1990-2017) of the means of the rate of CVD and risk factor exposures were averaged using the computer software program R.

To weigh the data according to population, internet searches (mostly Wikipedia) yielded the most recent population estimates for countries and subnational states, provinces, and regions. The 1120 GBD cohorts available were population weighted by a software program in R, resulting in an analysis dataset with 7846 population weighted cohorts representing about 7.8 billion people in 2020. Each male or female cohort in the population-weighted analysis dataset represented approximately 1 million people (range: < 100,000 to 1.5 million). World population data from the World Bank or the Organization for Economic Co-operation and Development could not be used because they did not include all 195 countries or any subnational data.

Supplementary Table 2 details how omega-3 fatty acid g/d was converted to fish g/d using data on the omega-3 fatty acid content of frequently eaten fish from the National Institutes of Health Office of Dietary Supplements (USA).^11^ As shown in Supplementary Table 3, we converted all of the animal and plant food data, including alcohol and sugary beverage consumption, from g/d to kilocalories/day (Kcal/day). For the g/d to Kcal/day conversions, we used the Nutritionix track app,^12^ which tracks types and quantities of foods consumed. Saturated fatty acids (SFA: 0-1 portion of the entire diet Kcal/day) was not available with GBD 2017 data, so GBD SFA risk factor data from GBD 2016 was used. Polyunsaturated fatty acid (PUFA) and trans fatty acid (TFA) GBD risk factor data from 2017 (0-1 portion of the entire diet Kcal/day) were also utilised, but monounsaturated fat data were not available. These fatty acid data expressed as 0-1 portion of the entire diet were converted to Kcal/day by multiplying by the total Kcal/day available (a covariate from the Food and Agriculture Organization^9^) per capita for each cohort.

### Outcome variable and covariates

CVD, the principal outcome variable of this analysis, was a combination variable consisting of the deaths/100k/year of male and female cohorts from (1) ischemic heart disease, (2) stroke, (3) hypertensive heart disease, (4) rheumatic heart disease, (5) non-rheumatic valvular disease, (6) subarachnoid haematoma, (7) myocarditis, (8) alcoholic cardiomyopathy, (9) endocarditis, aortic aneurysm, and (10) atrial fibrillation. Given the multiple dietary and other influences on the causation of CVD, we did not attempt to differentiate between dietary risk factors and covariates.

### Statistical methods

To determine the strengths of the risk factor correlations with CVD of population weighted worldwide cohorts (n=7846 cohorts) or subgroups of cohorts (e.g., highest and lowest CVD cohorts, etc.), we utilised Pearson correlation coefficients: r, 95% confidence intervals (CIs), and *p* values. To identify confounding in the univariate correlations, we used partial correlations, holding constant the suspected confounder variables.

### Sensitivity analyses

We performed subset analyses on the 1000 cohorts with the lowest and highest levels of CVD.

We used SAS and SAS OnDemand for Academics software 9.4 (SAS Institute, Cary, NC) for the data analysis.

## Results

Table 1 shows the 44 dietary, metabolic, and other worldwide risk factors potentially contributing to CVD (n=7846 cohorts). See Supplementary Table 1 for definitions of the risk factor and covariate variables. In Table 1, low density lipoprotein cholesterol mmol/L (LDL-C) negatively correlated with CVD worldwide (r=-0.279, 95% CI -0.299 to -0.258, p<0001,), meaning the higher the LDL-C the lower the CVD. Figure 1 gives the worldwide plot of LDL-C versus CVD, showing widely scattered data points with a negative slope of the least square regression line.

**Figure 1.**
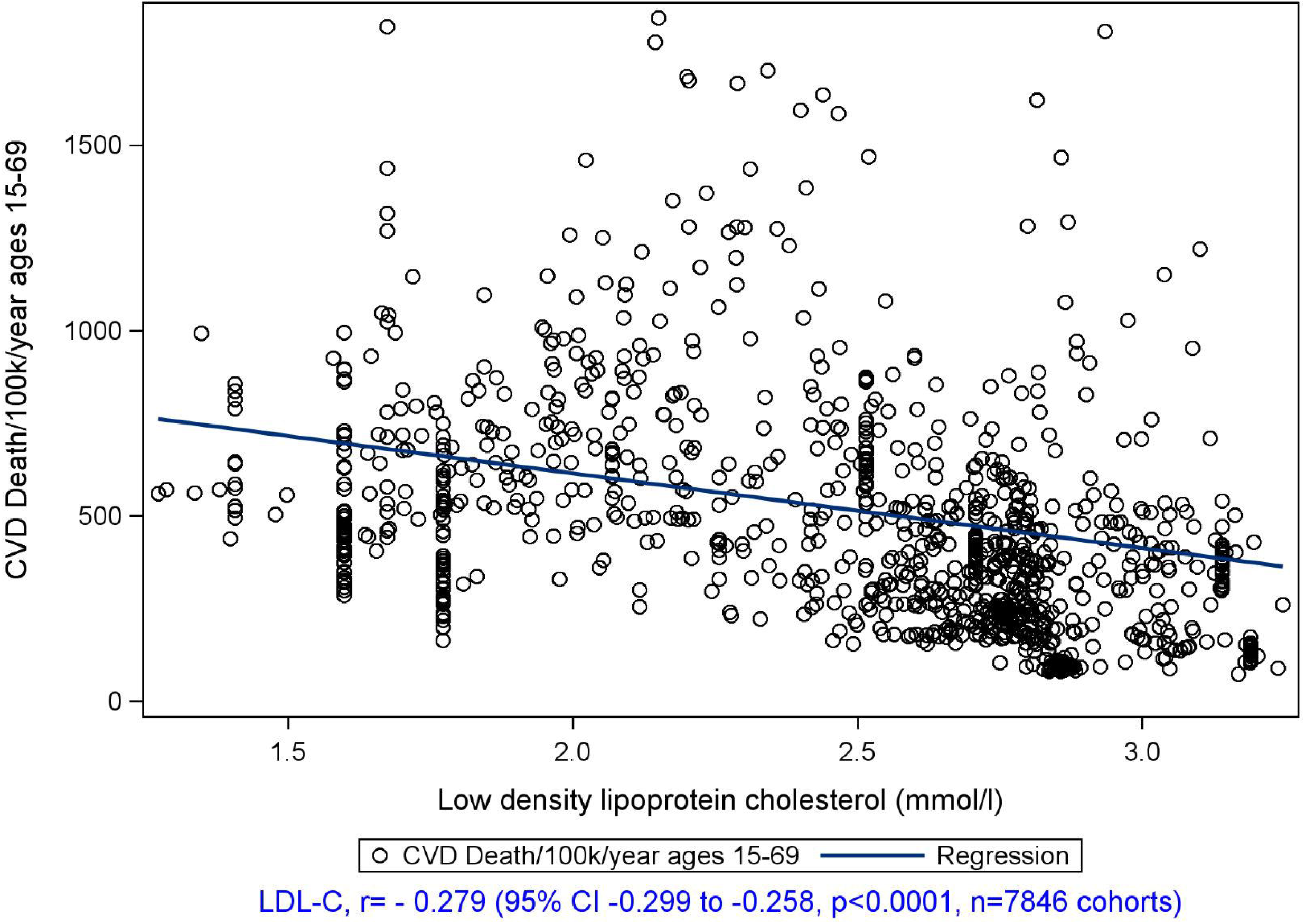

**Table 1.** Dietary and other risks related to CVD worldwide (n=7846 cohorts)

| CVD and CVD risk factors | Mean | SD | Min | Max | r | 95%<br>CI | 95%<br>CI | p |
| --- | --- | --- | --- | --- | --- | --- | --- | --- |
| Cardiovascular disease deaths/100k/year<br>ages 15-69 (CVD) | 543.66 | 288.01 | 73.47 | 1844 |  |  |  |  |
| CVD mean m/f | 543.69 | 246.13 | 135.46 | 1727 | 0.855 | 0.849 | 0.860 | <.0001 |
| LDL cholesterol mmol/L | 2.35 | 0.40 | 1.27 | 3.25 | -0.279 | -0.299 | -0.258 | <.0001 |
| Fat-soluble vitamin variable Kcal/day<br>(FSVV) | 285.36 | 193.31 | 58.78 | 932.18 | -0.329 | -0.349 | -0.309 | <.0001 |
| Processed meat Kcal/day | 5.33 | 9.72 | 0.20 | 68.77 | -0.204 | -0.226 | -0.183 | <.0001 |
| Red meat Kcal/day | 50.27 | 45.13 | 3.21 | 235.95 | -0.232 | -0.253 | -0.211 | <.0001 |
| Fish Kcal/day | 9.99 | 36.52 | 0.40 | 370.36 | -0.203 | -0.224 | -0.181 | <.0001 |
| Milk Kcal/day | 25.04 | 27.05 | 1.06 | 146.82 | -0.192 | -0.214 | -0.171 | <.0001 |
| Poultry Kcal/day available | 44.32 | 50.08 | 1.06 | 411.87 | -0.289 | -0.309 | -0.268 | <.0001 |
| Eggs Kcal/day available | 19.36 | 14.71 | 0.79 | 69.64 | -0.390 | -0.408 | -0.371 | <.0001 |
| Added Saturated fatty acids Kcal/day | 87.67 | 29.41 | 32.56 | 221.29 | -0.239 | -0.260 | -0.219 | <.0001 |
| Added PUFAs Kcal/day | 37.30 | 33.78 | 1.35 | 175.40 | -0.316 | -0.336 | -0.296 | <.0001 |
| Added Trans fatty acids Kcal/day | 6.09 | 6.28 | 0.91 | 35.77 | -0.104 | -0.126 | -0.082 | <.0001 |
| Alcohol Kcal/day | 81.03 | 57.33 | 4.25 | 429.81 | -0.061 | -0.083 | -0.039 | <.0001 |
| Sugary beverages Kcal/day | 298.36 | 152.38 | 72.91 | 1472.00 | 0.113 | 0.091 | 0.135 | <.0001 |
| Potatoes Kcal/day available | 84.04 | 74.60 | 3.07 | 533.88 | 0.050 | 0.028 | 0.073 | <.0001 |
| Corn Kcal/day available | 34.72 | 48.28 | 0.16 | 305.17 | -0.062 | -0.084 | -0.040 | <.0001 |
| Fruits Kcal/day | 40.21 | 22.50 | 3.58 | 161.39 | -0.355 | -0.374 | -0.336 | <.0001 |
| Vegetables Kcal/day | 79.76 | 43.12 | 9.48 | 304.17 | -0.107 | -0.128 | -0.085 | <.0001 |
| Nuts and seeds Kcal/day | 8.41 | 8.36 | 0.05 | 102.99 | -0.277 | -0.297 | -0.256 | <.0001 |
| Whole grains Kcal/day | 55.65 | 30.93 | 1.14 | 235.10 | -0.194 | -0.216 | -0.173 | <.0001 |
| Legumes Kcal/day | 51.74 | 32.23 | 0.51 | 194.70 | -0.024 | -0.046 | -0.002 | 0.0319 |
| Rice Kcal/day available | 141.86 | 116.34 | 1.42 | 461.80 | 0.007 | -0.015 | 0.029 | 0.5417 |
| Sweet potatoes Kcal/day available | 22.76 | 35.95 | 0.02 | 364.74 | -0.167 | -0.189 | -0.146 | <.0001 |
| Total Kcal/day available | 2574 | 418 | 1579 | 3898 | -0.203 | -0.224 | -0.181 | <.0001 |
| Vit A deficiency children/100k/year | 23205 | 10939 | 1267 | 50969 | 0.210 | 0.189 | 0.231 | <.0001 |
| Sodium g/d | 4.45 | 2.34 | 1.33 | 9.21 | -0.214 | -0.235 | -0.193 | <.0001 |
| Calcium g/d | 0.301 | 0.179 | 0.081 | 1.044 | -0.169 | -0.191 | -0.148 | <.0001 |
| Dietary fiber g/d | 9.21 | 3.15 | 2.72 | 22.68 | 0.019 | -0.003 | 0.041 | 0.0932 |
| Physical activity METs | 4714 | 1368 | 1609 | 7669 | 0.160 | 0.139 | 0.182 | <.0001 |
| Child underweight >2SD | 0.186 | 0.171 | 0.004 | 0.535 | 0.302 | 0.282 | 0.322 | <.0001 |
| Stop breast feeding <6 months | 0.119 | 0.055 | 0.016 | 0.242 | -0.302 | -0.322 | -0.282 | <.0001 |
| Ambient pollution PM<sub>0.25</sub> | 44.73 | 26.46 | 4.38 | 95.54 | 0.153 | 0.131 | 0.174 | <.0001 |
| Smoking rate (0-1) | 0.205 | 0.176 | 0.003 | 0.640 | 0.297 | 0.277 | 0.317 | <.0001 |
| Secondhand smoking (0-1) | 0.376 | 0.155 | 0.164 | 0.796 | -0.225 | -0.246 | -0.204 | <.0001 |
| Sublingual tobacco rate (0-1) | 0.068 | 0.095 | 0.001 | 0.419 | 0.284 | 0.264 | 0.304 | <.0001 |
| Blood lead level mcg/dl | 5.01 | 1.01 | 1.22 | 8.37 | 0.180 | 0.159 | 0.201 | <.0001 |
| Household air pollution (0-1) | 0.482 | 0.325 | 0.000 | 0.996 | 0.179 | 0.158 | 0.201 | <.0001 |
| Kidney disease stage III (0-1) | 0.056 | 0.028 | 0.015 | 0.154 | 0.194 | 0.173 | 0.215 | <.0001 |
| Type 1 DM early deaths | 10.37 | 9.39 | 0.55 | 112.49 | 0.340 | 0.320 | 0.359 | <.0001 |
| Type 2 DM early deaths | 17.50 | 15.65 | 0.63 | 269.67 | 0.227 | 0.205 | 0.247 | <.0001 |
| BMI kg/M<sup>2</sup> | 21.77 | 2.29 | 17.95 | 29.39 | -0.240 | -0.261 | -0.219 | <.0001 |
| Fasting plasma glucose mmol/L | 4.30 | 0.35 | 3.32 | 5.58 | -0.178 | -0.200 | -0.157 | <.0001 |
| Systolic BP mm Hg | 133.91 | 4.32 | 123.41 | 147.89 | 0.195 | 0.174 | 0.216 | <.0001 |
| Socio-demographic index (0-1) | 0.543 | 0.174 | 0.112 | 0.896 | -0.337 | -0.357 | -0.317 | <.0001 |
| Sex male 1 and female 2 | 1.50 | 0.50 | 1.00 | 2.00 | -0.395 | -0.414 | -0.376 | <.0001 |

Table 1 shows that CVD negatively correlated with all the animal foods and all the fatty acids (processed meats, red meats, fish, milk, poultry, eggs, SFA, PUFA, and TFA). Based on these unexpected finding, we formulated our “fat-soluble vitamin hypothesis”—insufficient dietary intake of fat-soluble vitamin containing animal foods and added fat for gut absorption increases cardiovascular disease risk. To test this hypothesis, we summed the Kcal/day of animal foods and added fats to produce a fat-soluble vitamins variable (Kcal/day) (FSVV =processed meats Kcal/day + red meats Kcal/day + fish Kcal/day + milk Kcal/day + poultry Kcal/day + eggs Kcal/day + added SFA Kcal/day + added PUFA Kcal/day + added TFA Kcal/day). In determining the portion of SFA, PUFA, and TFA added in addition to the fatty acids in the animal and plant foods, an adjustment factor (fatty acids * 0.46), adapted from the website “Our World in Data,” differentiated the fatty acids in individual foods (54% of the total) from the added fatty acids in oils, butter, lard, etc. (46% of the total).^13^ This adjustment prevented double counting fatty acids (Kcal/day) and allowed a more accurate determination of FSVV.

FSVV (with abundant SFA in animal foods and added extracted fat) correlated strongly with LDL-C worldwide (r=0.780, 95% CI 0.771 to 0.788, p<0001, n=7846 cohorts). Consequently, we considered that FSVV could serve as our marker for the lipid hypothesis as well as the fat-soluble vitamin hypothesis.

Table 2 shows a subgroup of 500 pairs of male and female cohorts with the lowest CVD values, totaling 1000 total cohorts (an arbitrary number). Notably, the mean FSVV in this low CVD subset (FSVV=531.38 Kcal/day) was much higher than the mean FSVV worldwide (FSVV=285.36 Kcal/day, Table 1) or the highest CVD subset mean FSVV (FSVV=235.66 Kcal/day, n=1000 cohorts, Table 2).

**Table 2.** CVD lowest (≈1 billion people) and highest 1000 cohorts (≈1 billion people)

| | CVD lowest $\approx 1$ billion people (n=1000 cohorts) | | | | CVD highest $\approx 1$ billion people (n=1000 cohorts) | | | |
| --- | --- | --- | --- | --- | --- | --- | --- | --- |
| CVD and risk factors | Mean | SD | Min | Max | Mean | SD | Min | Max |
| Cardiovascular Disease deaths/100k/year | 227.81 | 95.25 | 73.47 | 422.36 | 1045 | 356.90 | 430.20 | 1844 |
| CVD male/female mean | 227.81 | 42.52 | 135.46 | 292.97 | 1045 | 200.47 | 802.24 | 1727 |
| LDL cholesterol mmol/L | 2.808 | 0.256 | 1.598 | 3.247 | 2.376 | 0.380 | 1.35 | 3.20 |
| Fat-soluble vitamin variable Kcal/day (FSVV) | 531.4 | 180.20 | 143.63 | 912.69 | 235.66 | 99.68 | 80.53 | 503.98 |
| Processed meat Kcal/day | 15.30 | 12.20 | 0.61 | 61.09 | 3.70 | 4.95 | 0.22 | 18.12 |
| Red meat Kcal/day | 90.72 | 48.06 | 12.03 | 235.95 | 36.82 | 29.40 | 7.07 | 172.00 |
| Fish Kcal/day | 49.54 | 92.63 | 2.85 | 370.36 | 4.56 | 2.69 | 0.78 | 12.97 |
| Milk Kcal/day | 55.64 | 35.39 | 7.70 | 146.82 | 25.61 | 16.78 | 1.06 | 75.87 |
| Poultry Kcal/day available | 90.44 | 47.27 | 4.74 | 240.56 | 30.23 | 22.39 | 2.78 | 100.04 |
| Eggs Kcal/day available | 36.63 | 11.25 | 4.77 | 69.64 | 14.73 | 13.63 | 1.51 | 46.81 |
| Added Saturated fatty acids Kcal/day | 113.40 | 36.53 | 59.52 | 221.29 | 87.22 | 24.03 | 33.03 | 205.85 |
| Added PUFAs Kcal/day | 69.80 | 35.78 | 15.77 | 175.40 | 25.48 | 14.54 | 2.73 | 84.90 |
| Added Trans fatty acids Kcal/day | 9.91 | 9.25 | 1.65 | 34.90 | 7.31 | 8.62 | 1.21 | 35.77 |
| Alcohol Kcal/day | 119.72 | 66.67 | 4.25 | 296.70 | 45.83 | 46.24 | 5.77 | 340.37 |
| Sugary beverages Kcal/day | 320.29 | 251.94 | 72.91 | 1472 | 303.30 | 63.68 | 180.80 | 685.42 |
| Potatoes Kcal/day available | 81.34 | 51.14 | 9.50 | 177.30 | 98.34 | 96.77 | 3.94 | 533.88 |
| Corn Kcal/day available | 48.19 | 75.26 | 1.79 | 287.14 | 31.77 | 37.30 | 0.20 | 305.17 |
| Fruits Kcal/day | 64.84 | 16.49 | 23.05 | 128.61 | 31.23 | 13.93 | 3.58 | 70.11 |
| Vegetables Kcal/day | 108.02 | 44.60 | 9.48 | 214.26 | 83.83 | 56.67 | 14.43 | 198.51 |
| Nuts and seeds Kcal/day | 15.90 | 10.50 | 0.66 | 39.80 | 4.81 | 2.91 | 0.05 | 17.91 |
| Whole grains Kcal/day | 64.71 | 38.62 | 1.61 | 173.46 | 28.30 | 33.01 | 1.14 | 156.75 |
| Legumes Kcal/day | 47.88 | 25.99 | 2.95 | 133.26 | 31.55 | 18.93 | 0.51 | 103.17 |
| Rice Kcal/day available | 78.10 | 97.95 | 7.58 | 349.08 | 75.93 | 107.38 | 1.42 | 461.80 |
| Sweet potatoes Kcal/day available | 3.92 | 5.14 | 0.04 | 27.51 | 5.84 | 20.20 | 0.02 | 364.74 |
| Total Kcal/day available | 2972 | 409 | 1948 | 3572 | 2627 | 409 | 1579 | 3254 |
| Vit A deficiency children/100k/year | 14181 | 8723 | 1400 | 44100 | 20559 | 11395 | 1722 | 50969 |
| Sodium g/d | 3.92 | 1.22 | 1.33 | 6.70 | 3.30 | 0.76 | 1.33 | 5.96 |
| Calcium g/d | 0.506 | 0.189 | 0.183 | 1.044 | 0.321 | 0.150 | 0.089 | 0.792 |
| Dietary fiber g/d | 10.58 | 2.90 | 5.15 | 17.95 | 9.53 | 3.32 | 3.87 | 18.83 |
| Physical activity METs | 3802 | 1164 | 2162 | 7607 | 4273 | 1497 | 1838 | 7607 |
| Child underweight >2SD | 0.038 | 0.043 | 0.004 | 0.242 | 0.186 | 0.141 | 0.011 | 0.411 |
| Stop breast feeding <6 months | 0.176 | 0.032 | 0.071 | 0.218 | 0.116 | 0.044 | 0.036 | 0.208 |
| Ambient pollution PM<sub>0.25</sub> | 17.60 | 8.27 | 4.38 | 38.42 | 40.81 | 22.19 | 7.90 | 77.69 |
| Smoking rate (0-1) | 0.217 | 0.121 | 0.012 | 0.448 | 0.212 | 0.190 | 0.005 | 0.627 |
| Secondhand smoking (0-1) | 0.328 | 0.097 | 0.164 | 0.569 | 0.397 | 0.158 | 0.164 | 0.780 |
| Sublingual tobacco rate (0-1) | 0.010 | 0.015 | 0.001 | 0.111 | 0.060 | 0.072 | 0.001 | 0.238 |
| Blood lead level mcg/dl | 4.08 | 1.00 | 1.22 | 6.85 | 4.74 | 1.28 | 1.92 | 8.37 |
| Household air pollution (0-1) | 0.103 | 0.182 | 0.001 | 0.839 | 0.404 | 0.353 | 0.002 | 0.993 |
| Kidney disease stage III (0-1) | 0.042 | 0.020 | 0.015 | 0.111 | 0.078 | 0.024 | 0.035 | 0.139 |
| Type 1 DM early deaths | 4.79 | 3.88 | 0.59 | 20.65 | 16.38 | 15.31 | 2.62 | 112.49 |
| Type 2 DM early deaths | 15.69 | 18.23 | 0.75 | 87.92 | 23.65 | 24.88 | 0.89 | 269.67 |
| BMI kg/M<sup>2</sup> | 23.74 | 1.60 | 19.61 | 27.06 | 22.46 | 2.15 | 17.95 | 26.08 |
| Fasting plasma glucose mmol/L | 4.56 | 0.25 | 3.54 | 5.12 | 4.28 | 0.42 | 3.38 | 5.18 |
| Systolic BP mm Hg | 133.26 | 4.41 | 123.41 | 142.15 | 136.86 | 4.39 | 124.59 | 147.89 |
| Socio-demographic index (0-1) | 0.757 | 0.125 | 0.351 | 0.896 | 0.497 | 0.183 | 0.186 | 0.824 |
| Sex male 1 and female 2 | 1.50 | 0.50 | 1.00 | 2.00 | 1.50 | 0.50 | 1.00 | 2.00 |

To test the lipid hypothesis with FSVV as the marker variable, we calculated the median FSVV of the low CVD 1000 cohorts (median FSVV=567.27 Kcal/day) and formed a subset of the highest FSVV cohorts (FSVV≥567.27 Kcal/day, n=974 cohorts, Table 3). In Table 3, we see a much higher mean FSVV (FSVV≥692.58 Kcal/day) than in Table 2 and a strong positive correlation of FSVV with CVD (r=0.523, 95% CI 0.476 to 0.567, p<0001, n=974 cohorts). In contrast, LDL-C and CVD were negatively correlated in this high FSVV subset (r=-0.254, 95% CI -0.194 to -0.312, *p*<0001, n=974 cohorts). Figure 2 graphs the contrast between worldwide FSVV negatively correlating with CVD versus FSVV positively correlating with CVD in the highest FSVV cohorts (FSVV≥567.27 Kcal/day).

**Figure 2.**
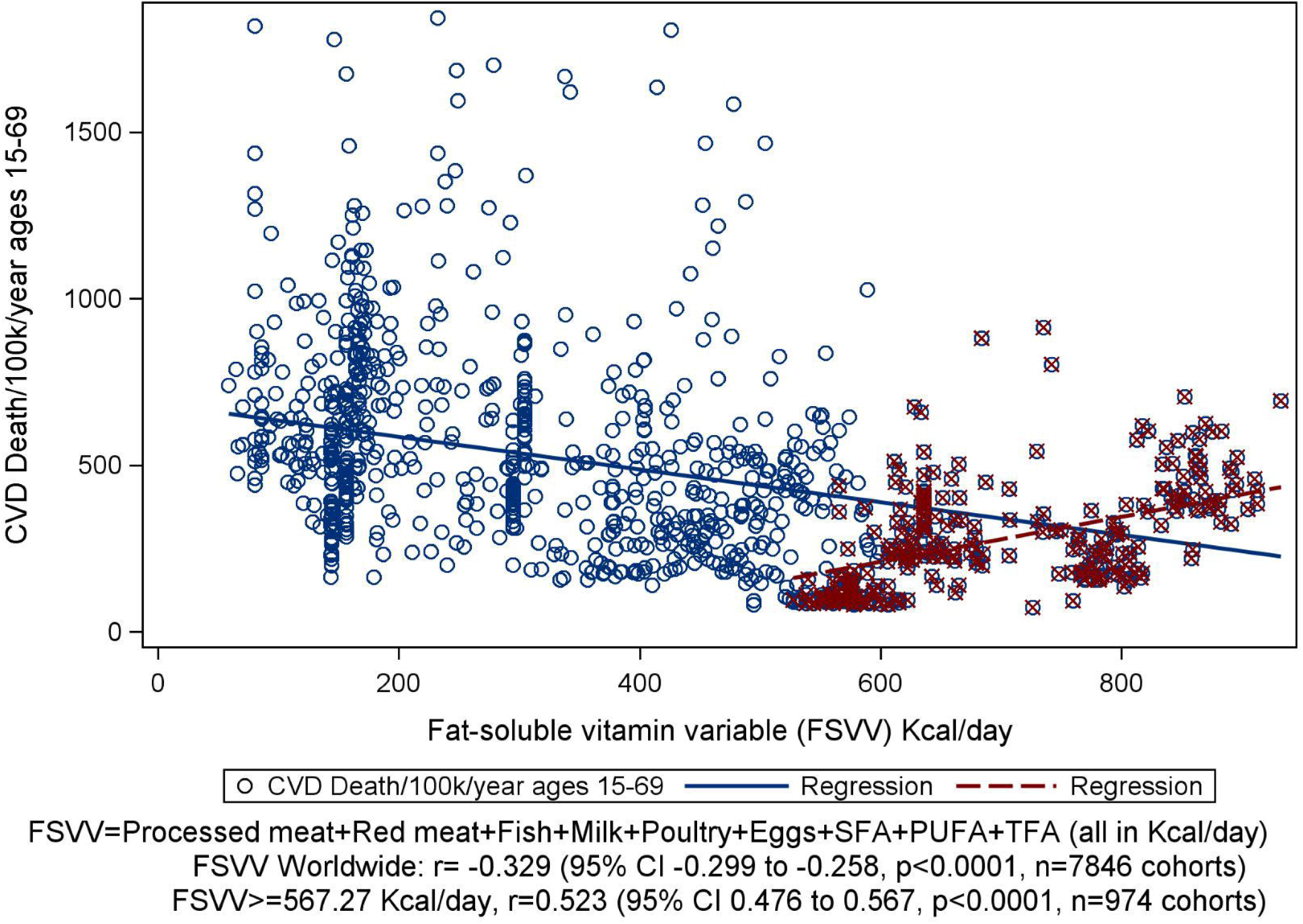

**Table 3.** CVD and risk factors for countries with FSVV 567.27 Kcal/day (n=974 cohorts)

| CVD and CVD risk factors | Mean | SD | Min | Max | r | 95%<br>CI low | 95% CI<br>high | P |
| --- | --- | --- | --- | --- | --- | --- | --- | --- |
| **Cardiovascular deaths/100k/year**<br>**ages 15-69** | 273.72 | 143.95 | 73.47 | 913.2 |  |  |  |  |
| **LDL cholesterol mmol/L** | 2.91 | 0.14 | 2.36 | 3.20 | -0.254 | -0.312 | -0.194 | <.0001 |
| **Fat-soluble vitamin variable** | 692.6 | 111.9 | 527.7 | 932.2 | 0.523 | 0.476 | 0.567 | <.0001 |
| **Processed meat Kcal/day** | 25.01 | 15.30 | 2.16 | 68.77 | 0.459 | 0.408 | 0.507 | <.0001 |
| **Red meat Kcal/day** | 122.2 | 41.41 | 43.65 | 236.0 | 0.655 | 0.618 | 0.690 | <.0001 |
| **Fish Kcal/day** | 49.08 | 89.24 | 7.60 | 370.4 | -0.259 | -0.317 | -0.199 | <.0001 |
| **Milk Kcal/day** | 78.58 | 26.14 | 14.60 | 146.8 | 0.185 | 0.123 | 0.245 | <.0001 |
| **Poultry Kcal/day available** | 130.0 | 60.21 | 41.65 | 411.9 | 0.366 | 0.311 | 0.419 | <.0001 |
| **Eggs Kcal/day available** | 40.80 | 7.25 | 14.59 | 69.64 | -0.242 | -0.301 | -0.182 | <.0001 |
| **Saturated fatty acids Kcal/day** | 299.9 | 66.83 | 129.4 | 481.0 | 0.193 | 0.132 | 0.253 | <.0001 |
| **PUFAs Kcal/day** | 211.9 | 83.06 | 80.53 | 381.3 | 0.382 | 0.327 | 0.435 | <.0001 |
| **Trans fatty acids Kcal/day** | 25.17 | 17.82 | 4.23 | 66.89 | 0.224 | 0.163 | 0.283 | <.0001 |
| **Alcohol Kcal/day** | 155.0 | 65.23 | 11.92 | 429.8 | 0.475 | 0.425 | 0.522 | <.0001 |
| **Sugary beverages Kcal/day** | 231.7 | 121.9 | 72.91 | 769.9 | 0.332 | 0.274 | 0.386 | <.0001 |
| **Potatoes Kcal/day available** | 106.4 | 40.47 | 9.50 | 225.0 | 0.109 | 0.046 | 0.170 | 0.00 |
| **Corn Kcal/day available** | 19.45 | 9.41 | 1.79 | 47.39 | 0.086 | 0.024 | 0.148 | 0.01 |
| **Fruits Kcal/day** | 66.91 | 15.93 | 34.83 | 161.4 | -0.014 | -0.077 | 0.049 | 0.66 |
| **Vegetables Kcal/day** | 117.4 | 34.24 | 42.27 | 304.2 | -0.127 | -0.188 | -0.064 | <.0001 |
| **Nuts and seeds Kcal/day** | 21.98 | 8.91 | 0.85 | 103.0 | 0.177 | 0.115 | 0.237 | <.0001 |
| **Whole grains Kcal/day** | 52.11 | 20.93 | 1.61 | 92.45 | 0.121 | 0.059 | 0.183 | 0.00 |
| **Legumes Kcal/day** | 40.76 | 25.29 | 2.95 | 133.3 | 0.040 | -0.023 | 0.102 | 0.21 |
| **Rice Kcal/day available** | 38.82 | 46.75 | 6.50 | 179.6 | -0.138 | -0.199 | -0.076 | <.0001 |
| **Sweet potatoes Kcal/day available** | 3.63 | 3.99 | 0.04 | 17.63 | -0.128 | -0.189 | -0.066 | <.0001 |
| **Total Kcal/day available** | 3219 | 292 | 2516 | 3898 | 0.249 | 0.189 | 0.307 | <.0001 |
| **Vit A deficiency children/100k/year** | 9316 | 6877 | 1400 | 28081 | -0.105 | -0.166 | -0.042 | 0.00 |
| **Sodium g/d** | 3.98 | 0.96 | 2.22 | 6.70 | -0.040 | -0.102 | 0.023 | 0.21 |
| **Calcium g/d** | 0.645 | 0.129 | 0.353 | 1.044 | 0.396 | 0.342 | 0.448 | <.0001 |
| **Dietary fiber g/d** | 10.62 | 1.75 | 6.30 | 18.15 | 0.241 | 0.181 | 0.299 | <.0001 |
| **Physical activity METs** | 3523 | 765 | 1781 | 5494 | 0.578 | 0.535 | 0.618 | <.0001 |
| **Child underweight >2SD** | 0.015 | 0.013 | 0.004 | 0.058 | -0.197 | -0.257 | -0.136 | <.0001 |
| **Stop breast feeding <6 months** | 0.196 | 0.017 | 0.129 | 0.219 | 0.268 | 0.208 | 0.325 | <.0001 |
| **Ambient pollution PM<sub>0.25</sub>** | 12.67 | 7.13 | 4.38 | 87.22 | 0.005 | -0.058 | 0.068 | 0.87 |
| **Smoking rate (0-1)** | 0.233 | 0.084 | 0.021 | 0.444 | 0.233 | 0.172 | 0.291 | <.0001 |
| **Secondhand smoking (0-1)** | 0.309 | 0.070 | 0.201 | 0.586 | -0.306 | -0.362 | -0.248 | <.0001 |
| **Sublingual tobacco rate (0-1)** | 0.013 | 0.020 | 0.001 | 0.125 | 0.579 | 0.536 | 0.620 | <.0001 |
| **Blood lead level mcg/dl** | 3.93 | 0.77 | 1.22 | 5.72 | 0.451 | 0.400 | 0.500 | <.0001 |
| **Household air pollution (0-1)** | 0.012 | 0.026 | 0.000 | 0.201 | -0.011 | -0.073 | 0.052 | 0.74 |
| **Kidney disease stage III (0-1)** | 0.036 | 0.012 | 0.016 | 0.109 | -0.028 | -0.090 | 0.035 | 0.39 |
| **Type 1 DM early deaths** | 3.93 | 2.67 | 0.55 | 15.97 | 0.573 | 0.529 | 0.613 | <.0001 |
| **Type 2 DM early deaths** | 9.54 | 7.52 | 0.75 | 54.22 | 0.622 | 0.582 | 0.659 | <.0001 |
| **BMI kg/M<sup>2</sup>** | 24.87 | 1.78 | 21.40 | 29.39 | 0.484 | 0.434 | 0.530 | <.0001 |
| **Fasting plasma glucose mmol/L** | 4.73 | 0.23 | 3.84 | 5.58 | 0.091 | 0.028 | 0.153 | 0.00 |
| **Systolic BP mm Hg** | 133.6 | 4.362 | 126.9 | 142.1 | -0.148 | -0.209 | -0.086 | <.0001 |
| **Socio-demographic index (0-1)** | 0.838 | 0.053 | 0.592 | 0.896 | -0.052 | -0.114 | 0.011 | 0.11 |
| **Sex male 1 and female 2** | 1.50 | 0.50 | 1.00 | 2.00 | -0.745 | -0.772 | -0.716 | <.0001 |

Table 4 shows FSVV’s component risk factors (processed meat, red meat, fish, milk, poultry, eggs, and added (SFA+PUFA+TFA)) for the 34 countries in Table 2 in descending order by CVD. Table 5 gives all the CVD risk factors for seven representative countries out of the 34 countries in Table 4 (CVD≤ 293.97, n=1000 cohorts).

**Table 4.** CVD related dietary risk factors (Kcal/day) in low CVD countries.

| **CVD lowest Countries (n=1000 cohorts)** | **n co-horts** | **CVD ascending order** | **FSVV** | **Processed meat** | **Red meat** | **Fish** | **Milk** | **Poultry** | **Eggs** | **Added SFA +PUFA +TFA** |
| --- | --- | --- | --- | --- | --- | --- | --- | --- | --- | --- |
| **Japan** | 158 | 169.19 | 618.69 | 19.19 | 58.72 | 260.54 | 29.04 | 63.93 | 52.23 | 135.04 |
| **France** | 64 | 174.00 | 644.78 | 13.59 | 135.07 | 11.17 | 95.75 | 99.86 | 43.30 | 246.04 |
| **Switzerland** | 8 | 175.04 | 558.41 | 11.50 | 129.01 | 6.75 | 110.90 | 55.57 | 31.20 | 213.47 |
| **Andorra** | 2 | 185.55 | 763.18 | 16.58 | 150.84 | 16.44 | 99.23 | 189.92 | 47.55 | 242.62 |
| **Peru** | 32 | 197.74 | 183.12 | 1.01 | 16.92 | 3.82 | 15.34 | 35.04 | 12.03 | 98.95 |
| **Spain** | 46 | 206.25 | 597.93 | 11.65 | 139.92 | 10.28 | 82.99 | 113.88 | 43.35 | 195.86 |
| **Italy** | 60 | 206.44 | 587.57 | 15.78 | 126.36 | 9.15 | 89.54 | 84.16 | 36.39 | 226.20 |
| **Iceland** | 2 | 206.51 | 609.62 | 11.97 | 112.73 | 11.67 | 85.37 | 63.57 | 29.51 | 294.81 |
| **Australia** | 24 | 208.03 | 698.26 | 9.79 | 164.14 | 14.15 | 97.05 | 153.33 | 24.92 | 234.89 |
| **Canada** | 36 | 222.11 | 624.98 | 25.31 | 126.28 | 18.48 | 76.13 | 147.46 | 34.94 | 196.39 |
| **Israel** | 8 | 228.32 | 662.00 | 5.95 | 59.50 | 15.35 | 68.79 | 240.56 | 40.54 | 231.30 |
| **South Korea** | 50 | 232.63 | 357.23 | 5.97 | 82.88 | 5.96 | 15.72 | 43.10 | 26.89 | 176.71 |
| **Belgium** | 12 | 235.98 | 636.39 | 12.28 | 122.36 | 11.19 | 93.44 | 85.00 | 33.28 | 278.86 |
| **Netherlands** | 18 | 236.61 | 629.56 | 15.72 | 133.06 | 8.66 | 114.16 | 72.70 | 41.67 | 243.59 |
| **Taiwan** | 24 | 237.37 | 587.36 | 3.01 | 106.58 | 9.15 | 15.95 | 132.38 | 34.87 | 285.41 |
| **Mexico** | 108 | 237.55 | 413.25 | 11.29 | 63.03 | 8.69 | 34.29 | 85.85 | 39.92 | 170.19 |
| **Panama** | 4 | 244.31 | 383.83 | 2.75 | 61.32 | 9.44 | 20.72 | 95.43 | 12.96 | 181.23 |
| **Sweden** | 10 | 247.21 | 563.83 | 29.31 | 119.77 | 11.80 | 124.13 | 45.42 | 35.04 | 198.37 |
| **New Zealand non Mauri** | 4 | 247.32 | 699.81 | 9.31 | 158.86 | 11.56 | 88.74 | 128.14 | 40.24 | 262.95 |
| **Norway** | 32 | 248.72 | 605.30 | 48.14 | 109.37 | 10.10 | 100.96 | 41.65 | 31.52 | 263.56 |
| **England** | 20 | 253.09 | 534.56 | 19.21 | 96.29 | 10.70 | 87.80 | 89.30 | 32.90 | 198.36 |
| **Costa Rica** | 4 | 260.73 | 408.42 | 2.63 | 51.74 | 8.12 | 46.55 | 78.05 | 28.17 | 193.14 |
| **Luxembourg** | 2 | 262.53 | 578.00 | 14.65 | 163.17 | 11.79 | 91.99 | 71.48 | 27.60 | 197.32 |
| **Chile** | 18 | 267.50 | 404.12 | 19.11 | 90.33 | 9.65 | 31.93 | 97.29 | 19.70 | 136.10 |
| **Guatemala** | 16 | 269.85 | 231.05 | 1.89 | 20.05 | 5.28 | 13.32 | 53.94 | 30.46 | 106.12 |
| **Kenya** | 12 | 270.04 | 150.04 | 1.41 | 25.83 | 3.09 | 18.89 | 4.74 | 4.77 | 91.31 |
| **Ecuador** | 16 | 270.64 | 375.35 | 6.87 | 56.88 | 7.34 | 35.51 | 59.92 | 14.66 | 194.16 |
| **USA** | 94 | 274.00 | 825.54 | 36.45 | 136.35 | 15.79 | 90.90 | 190.90 | 41.10 | 314.05 |
| **Denmark** | 6 | 275.10 | 600.48 | 14.46 | 137.75 | 13.38 | 98.01 | 74.42 | 45.96 | 216.51 |
| **Austria** | 8 | 277.67 | 621.00 | 8.95 | 194.65 | 9.26 | 98.68 | 70.00 | 39.96 | 199.50 |
| **Portugal** | 10 | 278.50 | 524.81 | 5.16 | 124.77 | 8.77 | 79.43 | 94.26 | 24.18 | 188.25 |
| **Nicaragua** | 6 | 282.04 | 214.24 | 1.52 | 14.93 | 4.07 | 20.66 | 41.76 | 17.41 | 113.90 |
| **Tehran (Iran)** | 16 | 283.29 | 299.65 | 1.23 | 22.63 | 5.36 | 19.23 | 65.90 | 18.89 | 166.41 |
| **Thailand** | 70 | 289.91 | 247.43 | 0.70 | 32.01 | 6.95 | 7.85 | 57.07 | 31.10 | 111.75 |
| **Total cohorts** | 1000 |  |  |  |  |  |  |  |  |  |

**Table 5.**
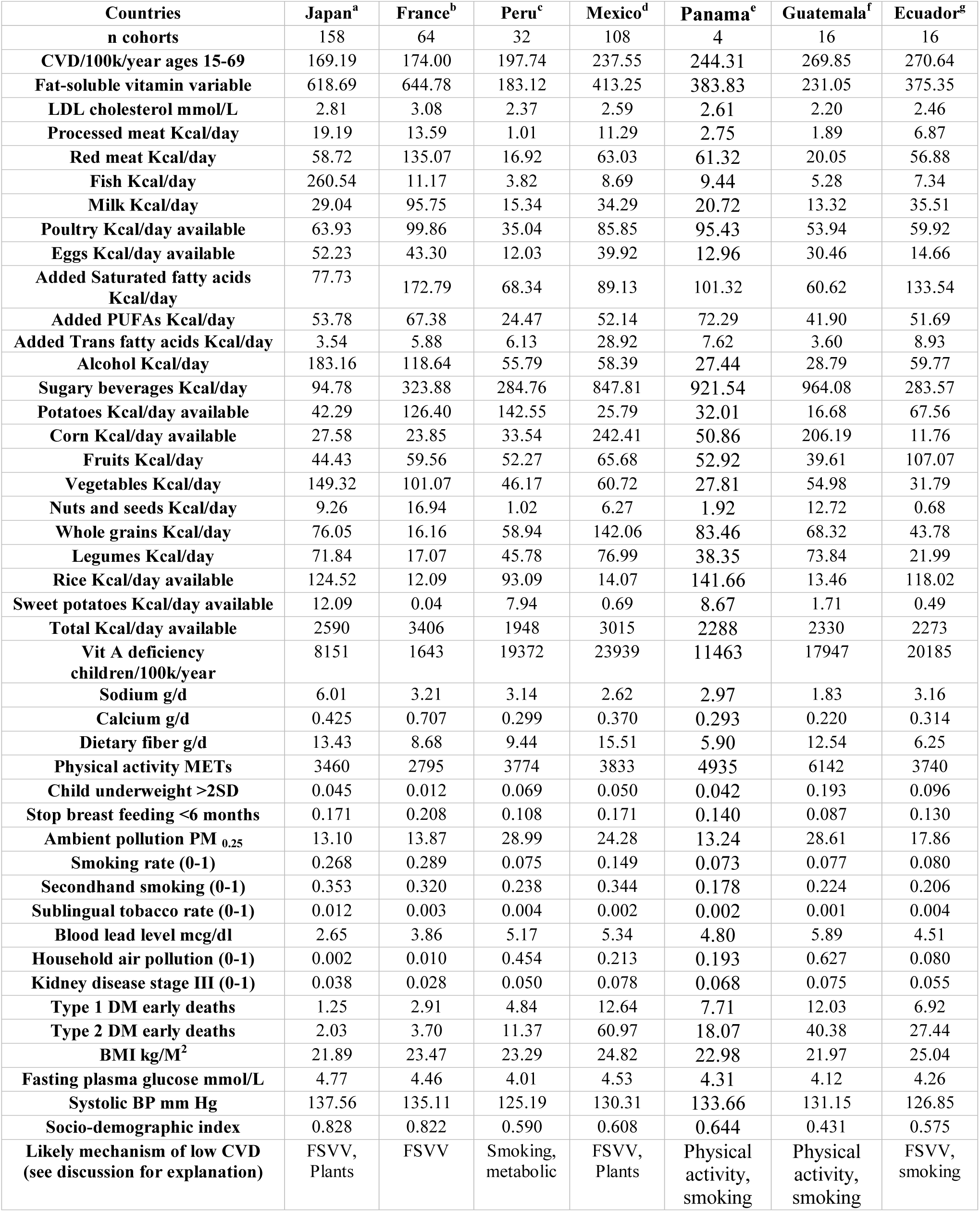
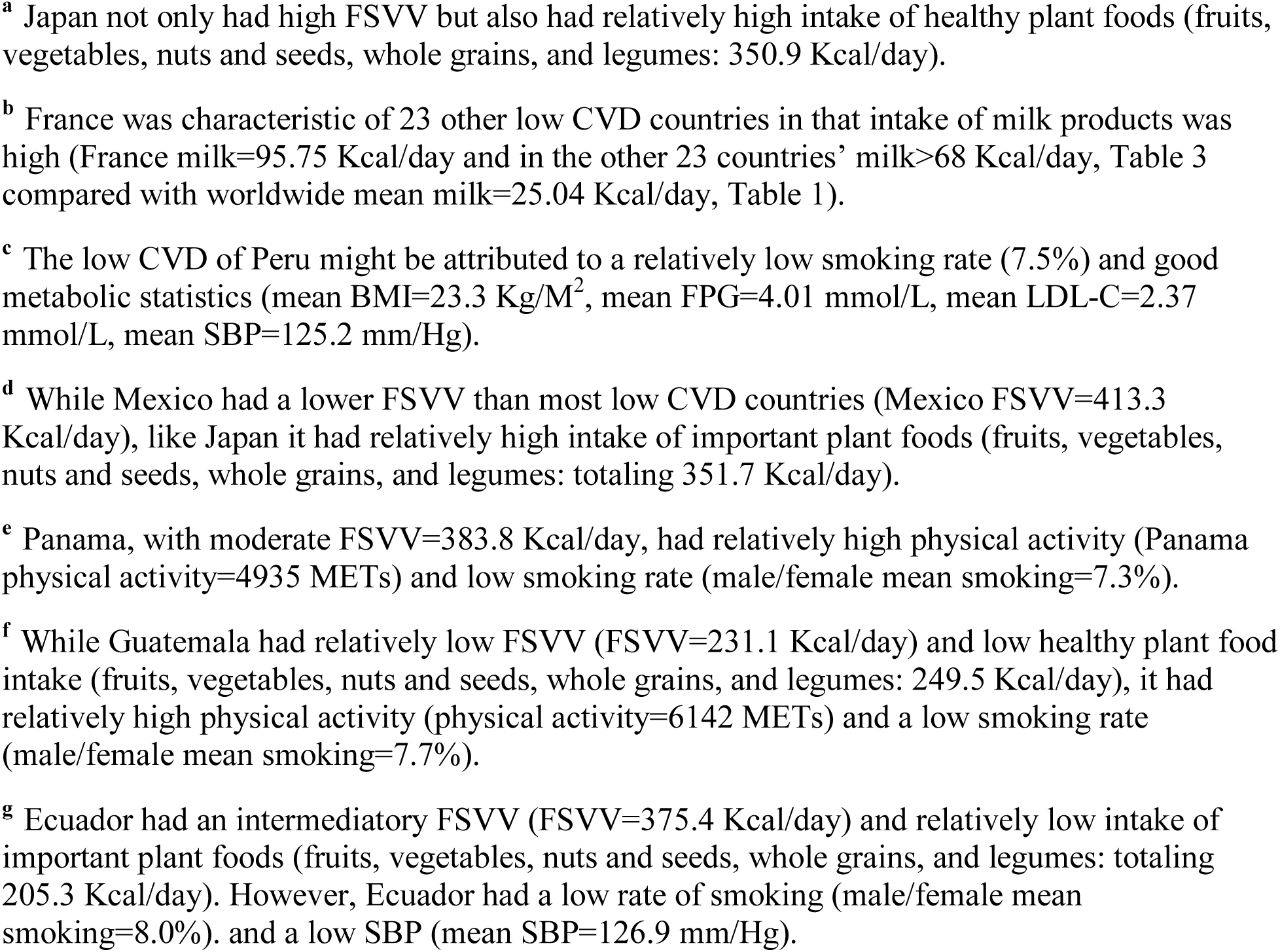
Representative examples of low CVD countries including all risk factors.

The average alcohol intake was 81.0 Kcal/day worldwide. Alcohol Kcal/day correlated negatively with CVD worldwide (r=-0.061, 95% CI -0.083 to -0.039, p<0001, Table 1). However, in high FSVV cohorts (FSVV≥567.27 Kcal/day), mean alcohol=155.02 Kcal/day), alcohol positively correlated with CVD (r=0.475, 95% CI 0.425 to 0.522, p<0001, n=974 cohorts, Table 3).

Vitamin A deficiency incidence/100k in children ≤ 5 years old, the only fat-soluble vitamin in the GBD database, correlated positively with CVD worldwide (r=0.210, 95% CI 0.189 to 0.231, p<0001, Table 1).

Dietary fiber was not significantly correlated with CVD worldwide (r=0.019, 95% CI -0.003 to 0.041, p=0.09, Table 1). Once dietary fiber was adjusted for FSVV, alcohol, sugary beverages, and potatoes by partial correlation analysis; dietary fiber correlated negatively with CVD (r=- 0.052, 95% CI -0.074 to -0.030, p<0.0001).

Physical activity positively correlated with CVD worldwide (r=0.160, 95% CI 0.139 to 0.182, p<0.0001, Table 1). However, confounding factors existed:

1. countries with higher FSVV levels also had less physical activity (FSVV versus physical activity worldwide: r=-0.366, 95% CI: -0.385 to -0.347, *p*< 0.0001),
2. females had less physical activity than males (mean physical activity males=5061 METs/week, mean physical activity females=4356 METs/week), along with less CVD (mean CVD males=656.2, mean CVD females=428.6), and
3. males smoked tobacco more than females (mean smoking (0-1) males=0.339 or 33.9%, mean smoking (0-1) females=0.070 or 7%), and males had more physical activity (as #2 above).

Once physical activity was adjusted for FSVV, smoking, and sex by partial correlation analysis, it correlated negatively with CVD (r=-0.127, 95% CI -0.149 to -0.104, p<0.0001).

In cohorts mostly in developed countries, with FSVV ≥567.27 Kcal/day, discontinuing breast feeding correlated positively with CVD (r=0.268, 95% CI 0.208 to 0.325, p<0.0001, Table 3).

Secondhand smoking negatively correlated with CVD worldwide (r=-0.225, 95% CI -0.246 to - 0.204, p<0.0001, Table 1). After adjusting secondhand smoking for (1) smoking, (2) sublingual tobacco use, (3) household smoke, (4) ambient air pollution, and (5) sex by partial correlation analysis; secondhand smoke correlated positively with CVD (r=0.048, 95% CI 0.025 to 0.070, p<0.0001).

The metabolic risk factors, BMI, FPG, and LDL-C, all correlated negatively with CVD worldwide (BMI r=-0.240, 95% CI -0.261 to -0.219, p<0.0001, FPG r=-0.178, 95% CI -0.200 to -0.157, p<0.0001, and LDL-C r=-0.279, 95% CI -0.299 to -0.258, p<0.0001, respectively, Table 1). Once they were adjusted for FSVV, alcohol, sugary beverages, and potatoes by partial correlation analysis; they all correlated positively with CVD (BMI r=0.028, 95% CI 0.006 to 0.050, p=0.0144, FPG r=0.076, 95% CI 0.054 to 0.098, p<0.0001, LDL-C r=0.170, 95% CI 0.148 to 0.191, p<0.0001, respectively). In the cohorts with FSVV≥567.27 Kcal/day, BMI and FPG positively correlated with CVD (BMI r=0.484, 95% CI 0.434 to 0.530, p<0.0001 and FPG r=0.091, 95% CI 0.028 to 0.153, p=0.0046, respectively).

## Discussion

Since this GBD data-based worldwide risk factor—health outcome analysis methodology has no precedent, there is no medical literature of previous such studies. The best comparator for this CVD versus risk factor analysis with IHME GBD data would be the comparative risk assessment (CRA) systematic literature review-based methodology used in drafting the “Planetary Health Diet.” Proposed by the EAT-Lancet Commission, the Planetary Health Diet was meant to spark a global transformation in human diets to improve health and mitigate climate change. The EAT-Lancet Commission authors said, “Transformation to healthy diets by 2050 will require substantial dietary shifts, including a greater than 50% reduction in global consumption of unhealthy foods such as red meat and sugar, and a greater than 100% increase in the consumption of healthy foods such as nuts, fruits, vegetables and legumes.”^14^ Reducing diet-caused non-communicable diseases (e.g., obesity, type 2 diabetes, CVD, cancers, etc.) was a major goal. WHO data show that about 44% of worldwide non-communicable disease deaths were CVD related,^15^ so these CVD analysis findings from GBD data can be fairly contrasted with the Planetary Health Diet proposal from CRA systematic literature review and expert opinion methodology.

When introduced by the EAT-Lancet Commission, the Planetary Health Diet sparked controversy. Dr. Francisco Zagmutt and colleagues disputed the rigor of the analytics, “A truly effective global solution to the problem of human nutrition and environmental impact must be replicable, transparent, and supported with correct quantification of its impact. Unfortunately, the report did not meet these criteria.”^16^ The World Health Organization (WHO) withdrew sponsorship over the exclusivity of one dietary approach for the entire planet and the negative impact of reducing cattle and pig ranching in developing countries.^17^

The methodologies and findings of this GBD data-based analysis of diet and other risk factors related to global CVD were markedly different than those of the EAT-Lancet Commission. Some of the consequential differences were as follows:

1. The Planetary Health Diet recommendations were based on systematic literature reviews primarily from researchers and participants in developed countries, largely missing at least 70% of the world’s population. This IHME GBD data-based population weighted analysis proportionally included people in developing countries.
2. To Dr. Zagmutt’s point about the lack of rigor of the Planetary Health Diet analytics, this IHME GBD data analysis used a single population weighted analysis dataset, spanning 28 years, making it replicable (with 2020 GBD data), transparent, and supported with reasonable quantification of the impact of global human nutrition on CVD.
3. The overall amounts of animal foods (Kcal/day) recommended by the Planetary Health Diet (mean animal foods: 304 Kcal/day)^18^ do not differ substantially from the overall animal foods (Kcal/day) in the GBD lowest CVD cohorts (338 Kcal/day, n=1000 cohorts, CVD < 292.97 deaths/year) in this analysis (Table 2). However, the Planetary Health Diet recommended 40 Kcal/day of fish^18^ while the GBD data show that the world fish consumption from 1990-2017 averaged 10.0 Kcal/day (Table 1). Levels of overfishing are unsustainable now, so quadrupling fish consumption worldwide is impossible. Likewise, the Planetary Health Diet recommended average milk consumption to be 153 Kcal/day^18^ while the GBD data show the world average milk consumption per person from 1990-2017 of 25.0 Kcal/day (Table 1). Increasing the world’s dairy cows six-fold, most of whom would eventually be slaughtered for meat, would seem to contradict major goals of the EAT-Lancet Commission—reducing methane emissions from cows and reducing red meat consumption. Instead, increasing all livestock production in developing countries would be projected to improve CVD outcomes (Table 1) as would decreasing cattle, pig, and poultry raising, but not fish or eggs, in high FSVV cohorts (Table 3).
4. Whereas the Planetary Health Diet has a single set of recommended ranges of quantities of plant and animal foods for all humanity, the GBD data-based analysis methodology can apply to people in developing countries as well as developed countries.

Fat-soluble vitamins include vitamins A, D, E, and K (K1=phylloquinones and K2= menaquinones). Vitamin K2 (menaquinones from animal foods and fermented plant products) inhibits arterial calcification, a major factor in the pathogenesis of atherosclerosis and cardiovascular disease.^19^ Vitamin A and its beta-carotene precursor are important in preventing congenital heart disease.^20, 21^ Vitamin E protects the vasculature against oxidative stress, including lipid peroxidation and the production of atherogenic forms of LDL-C—a part of the pathogenesis of cardiovascular disease.^22^ Regarding vitamin D, a meta-analysis of prospective studies showed an inverse correlation between cardiovascular disease and circulating 25(OH)-vitamin D ranging from 20 to 60 nmol/L.^23^

The four fat-soluble vitamins come primarily from animal foods. Vitamin K2 levels are high in aged cheeses, other fermented dairy products,^24^ organ meats (e.g., bovine and pork liver), chicken, and egg yolks.^25^ The Japanese had the world’s lowest CVD from 1990-2017 (mean CVD=169.2 deaths/100k/year). Paradoxically, crucial CVD risk factors were high in the Japanese (e.g., smoking: mean male/female smoking=26.8%, salt intake (mean sodium= 6.01 g/d, systolic blood pressure (mean SBP= 137.6 mm/Hg, Table 5). Relative to fermented cheeses, the content of vitamin K2 in fish was low to negligible (Japanese fish consumption mean=260.5 Kcal/day, Table 4).^25^ Instead, Japanese consumed high quantities of vitamin K2 in the form of fermented soy products like natto and miso.^26, 27^ People that also consumed high amounts of fermented legumes were the Taiwanese (soybean Douchi, soybean Meitauza, and soybean curd^28^), South Koreans (Cheonggukjang, a fermented soybean stew^28^), and Thais (soybean Thua nao^29^). The Taiwanese, South Koreans, and Thais were also listed among the 34 low CVD countries (Table 4).

In high FSVV cohorts, (FSVV≥ 567.27 Kcal/day), why LDL-C correlated negatively with FSVV (r=-0.254 95% CI -0.312 to -0.194, *p*<0.0001, n=974 cohorts) is not clear. However, multiple studies have documented that LDL-C may be influenced by genetic, racial, or ethnic differences that do not necessarily correspond with risk of CVD.^30, 31^ Additionally, LDL-C lowering medications in wide use especially in high FSVV intake developed countries may also confound the relationship of LDL-C with both LSVV and CVD risk in this subset cohort analysis.

Since there are high FSVV and low FSVV intake individual people widely distributed worldwide, prospective observational studies of individuals in high FSVV areas, rather than GBD cohort studies like this one, could better demonstrate positive correlations between LDL-C (or total cholesterol) and CVD. This may explain why the Seven Countries Study^32^ of Ancel Keys and the Framingham Heart Study,^33^ both in high FSVV regions, supported the lipid hypothesis. However, the Prospective Urban Rural Epidemiology (PURE) Study, with individual data on 135,335 patients from three high-income, 11 middle-income, and four low-income countries was clearly at variance with the lipid hypothesis.^34^ The PURE Study found that higher saturated fat intake was associated with lower risk of stroke (quintile 5 *vs* quintile 1, HR 0.79, (95% CI 0.64 to 0.98), p_trend_=0.0498).

Defining the optimal FSVV range that will reduce CVD risk while not increasing other diseases (e.g., cancers and obesity) will require more research. Supplements of fat-soluble vitamins to reduce the animal food and added fat intake required to minimise CVD should be studied, especially in developing countries with less access to animal foods.

Given the negative correlations of CVD with both fish and eggs in both tables 1 and 3, the reduction of FSVV in high FSVV countries might best come from meat, poultry, and added fats. Although milk (including other dairy products) correlated positively with CVD in Table 3 (r=0.185, 95% CI 0.123 to 0.245, p<0001), this probably related to Japan and Peru having the lowest and fifth lowest CVD (Japan: mean CVD=169.2, mean milk=29.0 Kcal/day, Peru: mean CVD=197.7, mean milk=15.3 Kcal/day). Without Japan and Peru in the analysis (n=834 cohorts), milk correlated negatively with CVD (r=-0.071, 95% CI -0.138 to -0.030, p=0.0405). Of the 34 low CVD countries in Table 4, 20 had high milk intakes (milk Kcal/day range: 68.8 Kcal/day-124.1Kcal/day versus mean milk worldwide=25.0 Kcal/day, Table 1) and these high milk intake cohorts all had relatively high per capita intakes of cheese.^35, 36^ Fermented, full-fat dairy products have high levels of fat-soluble vitamin K2,^24, 25^ which have been found to be important in preventing calcification of arteries and atherosclerosis.^37^

Potatoes Kcal/day available positively correlated with CVD both worldwide (r=0.050, 95% CI 0.028 to 0.073, p< 0.0001, Table 1) and in cohorts with FSVV≥ 567.27 Kcal/day (r=0.109, 95% CI 0.046 to 0.170, p< 0.0001, Table 3). Notably, half, or more of the potatoes consumed worldwide are in the form of highly processed food products.^38^ Recent large prospective observational studies have found higher consumption of ultra-processed foods associated with an increased risk of cardiovascular disease incidence and CVD mortality.^39, 40^ Data from 79 high- and middle-income countries showed that ultra-processed products dominate the food supplies of high-income countries and that their consumption is now rapidly increasing in middle-income countries.^41^

Dietary sodium negatively correlated with CVD worldwide (r=-0.213, 95% CI -0.234 to -0.192, p<0.0001, Table 1) and was not significantly correlated with CVD in high FSVV cohorts (FSVV≥ 567.27 Kcal/day: r=-0.040, 95% CI 0.023 to 0.170, p=0.21, Table 3) This was contrary to an analysis of sodium intake versus cardiovascular deaths in 66 countries by Mozaffarian, et. al who attributed 9.5% of cardiovascular deaths worldwide to high sodium intake.^42^ However, O’Donnell and colleagues found the relationship of sodium to cardiovascular disease to be J shaped curve and suggested that the lowest cardiovascular risk is with moderate sodium intake in the 3-5 g/d range.^43^ This analysis is consistent with O’Donnell’s study.

The moderately strong positive correlation of child ≤5 years old severe underweight (weight >2 SD below the mean for height) with CVD worldwide (r=0.306, 95% CI 0.285 to 0.325, p<0.0001, Table 1) suggested a relationship of infant/child malnutrition with later CVD. Babies surviving the so called “Dutch famine” toward the end of the second world war (1944-45) have been shown to have higher subsequent heart disease incidence than earlier or later cohorts in Holland.^44^ This suggested that in utero and early infancy severe malnutrition may subsequently contribute substantially to CVD, especially in developing countries.

Prematurely stopping breast feeding positively correlated with CVD in the countries with FSVV ≥567.27 Kcal/day (r=0.268 95% CI: 0.208 to 0.325, *p*=0.0027, Table 3). A review of observational studies of breast feeding related to metabolic risk factors for CVD in developed countries suggested that breast feeding was associated with increased insulin sensitivity and decreased systolic blood pressure in later life. Breast feeding also had metabolic benefits for the mother.^45^

According to the WHO, ambient air pollution (particulate matter ≤2.5 micrometers diameter (PM _2.5_)) causes cardiovascular and respiratory diseases and cancers.^46^ The WHO considers >10 μg/m^3^ of PM_2.5_ particles a health hazard. In this GBD data analysis, air pollution (PM_2.5_) in countries with FSVV≥567.27 Kcal/day and mean CVD=273.7 deaths/100k/year averaged 12.7 μg/m^3^. Worldwide, comparable numbers for air pollution (PM _2·5_) were mean=44.7 μg/m^3^ and mean CVD=543.7 (Table 1).

These IHME GBD data support the link of smoking tobacco to CVD (r=0.298, 95% CI 0.278 to 0.318, p<0.0001, Table 1). The mean incidence of smoking worldwide=20.5% (Table 1). In the 34 countries with the lowest CVD, the mean smoking incidence=23.3% (n=974 cohorts, Table 3). Smoking was clearly a major risk factor for CVD, but these data suggest that diet was more influential. Given the multiple dietary and other contributors to CVD, the CVD population attributable fraction (PAF) for smoking was probably far below the WHO estimate that tobacco accounts for 20% of coronary artery disease mortality.^47^

The positive correlation of blood lead levels with CVD worldwide (r=0.180, 95% CI 0.159 to 0.201, *p*<0.0001, Table 1) was consistent with reports that lead toxicity leads to hypertension.^48^ Also, environmental lead exposure has been linked with all-cause mortality, cardiovascular disease mortality, and ischaemic heart disease mortality in the US.^49^

This analysis showed that kidney disease correlated positively with CVD (r=0.194, 95% CI 0.173 to 0.215, p<0.0001, Table 1). A systematic literature review of kidney disease revealed that chronic kidney disease resulted in an estimated 1.2 million deaths in 2017, of which a large portion were from CVD.^50^ Other CVD risk factors may also lead to kidney disease (e.g., systolic hypertension, types 1 and 2 diabetes).

### Limitations

Our study was subject to all the limitations discussed in previous GBD publications.^51, 52^ These included gaps, biases, and inconsistencies in data sources as well as limitations in the methods of data processing and estimation. Having comprehensive data on dietary inputs is key to more accurate and reliable analyses. These GBD data on animal foods, plant foods, alcohol, sugary beverages, and fatty acids were not comprehensive and comprised only 1191.4 Kcal/day per person on average worldwide. Subnational data on all risk factors were available on only four countries. Because the data formatting and statistical methodology were new, this was necessarily a post hoc analysis and no pre-analysis protocol was possible. This GBD data analysis should be repeated with the most recently released GBD 2019 data when those data become available to volunteer collaborators.

### Generalisability

This new the GBD statistical analysis methodology for finding correlations between dietary and other risk factors and health outcomes (e.g., CVD) can be applied to any of the risk factors and health outcomes available with the new IHME GBD 2020 data. With further experience with the methodology, we can determine the generalisability.

## Conclusion

The lipid hypothesis (e.g., dietary SFA from animal foods and added fats causes CVD) was consistent with the strong positive correlation of FSVV with CVD in high FSVV cohorts. However, the lipid hypothesis was not supported worldwide. Global data analysis supported the fat-soluble vitamin hypothesis because FSVV negatively correlated with CVD worldwide. This methodology of analysing IHME GBD data might well have advantages over the CRA systematic literature review and expert opinion method in developing policy recommendations, clinical practice guidelines, and public health recommendations. The EAT-Lancet Commission might consider using IHME GBD data to guide another version of a planetary health diet.

## Data Availability

Our dataset, SAS codes, and Excel files are posted on the Mendeley data repository.

https://data.mendeley.com/datasets/g6b39zxck4/6

## Article information

### Contributors

DKC had full access to the GBD data in the study and takes responsibility for the integrity of the data and the accuracy of the data analysis. DKC conceived and designed the study, acquired and analysed the GBD data from IHME, interpreted the study findings, drafted the manuscript, critically reviewed and edited the manuscript and tables, and approved the final version for publication.

CW designed software programs in R to format and population weight the data, aided with the SAS statistical analysis, critically reviewed the manuscript, and approved the final version for publication.

### Data Sharing Statement

The formatted analysis dataset for this analysis, SAS codes, and Excel files are posted on the Mendeley data repository https://data.mendeley.com/datasets/g6b39zxck4/6)

### Additional Contributions

We thank Scott Glenn and Brent Bell from IHME who supplied us with the GBD risk factor exposure data.

### Checklist

This report followed the STrengthening the Reporting of OBservational studies in Epidemiology (STROBE) guidelines for reporting global health estimates.^53^

---

**Supplementary Table 1.** Definitions of GBD risk factors and covariates related to CVD.

| **Variables** | **Definition** |
| --- | --- |
| **Alcohol** | Any alcohol consumption (g/day) |
| **Ambient particulate matter pollution** | Annual average daily exposure to outdoor air concentrations of particulate matter with an aerodynamic diameter of $\leq 2.5 \mu\text{g}/\text{m}^3$ (PM<sub>2.5</sub>) |
| **Body-mass index** | Body mass index (BMI) ($\text{kg}/\text{m}^2$)—the dependent variable of interest |
| **Chewing tobacco** | Current use of any chewing tobacco product |
| **Child underweight** | Proportion of children – 3 SD to – 2 SD of the WHO 2006 standard weight-for-age curve (0-1) |
| **Corn** | Corn availability per capita (g/day), a covariate |
| **Discontinued breast feeding** | Proportion of children aged 6-23 months who do not receive any breast milk |
| **Eggs** | Eggs availability per capita (g/day) a covariate |
| **Fasting plasma glucose** | Fasting plasma glucose (mmol/L) |
| **Fish** | This variable expressed in g/day was derived by determining the weight of fish in g corresponding to 1 g of omega-3 fatty acids (eicosapentaenoic acid and docosahexaenoic acid) by averaging the fish g per 1 g of omega-3 fatty acids 20 species of fish= 117.04 g/day fish/1 g/day omega-3 fatty acids (Supplementary Table 3) |
| **Fruits** | Consumption of fruits (includes fresh, frozen, cooked, canned, or dried fruit but excludes fruit juices and salted or pickled fruits) (g/day) |
| **Household air pollution from solid fuels** | Individual exposure to PM<sub>2.5</sub> due to use of solid cooking fuel |
| **Kidney function impaired** | Proportion of the population with ACR $>30 \text{ mg}/\text{g}$ or GFR $<60 \text{ mL}/\text{min}/1.73 \text{ m}^2$, excluding end-stage renal disease |
| **Kilocalories available /day** | The mean number of kilocalories per capita available per day to people in each location (kcal/day available), a covariate |
| **LDL cholesterol** | Serum low-density lipoprotein cholesterol (mmol/L) |
| **Lead exposure** | Blood lead levels in $\mu\text{g}/\text{dL}$ of blood, bone lead levels in $\mu\text{g}/\text{g}$ of bone |
| **Legumes** | Consumption of beans, lentils, pulses (g/day) |
| **Milk** | Consumption of milk including non-fat, low-fat, and full-fat milk but excluding soy milk and other plant derivatives (g/day) |
| **Nuts and seeds** | Consumption of nuts and seeds (g/day) |
| **Physical activity** | Average weekly physical activity at work, home, transport-related and recreational measured by MET min per week. Less than 3000 METs per week constitutes low physical activity. |
| **Poultry** | Poultry availability per capita (g/day), a covariate |
| **Potatoes** | Potatoes availability per capita (g/day), a covariate |
| **Processed meat** | Consumption of any processed meat (includes meat preserved by smoking, curing, salting, or addition of chemical preservatives, including bacon, salami, sausages, or deli or luncheon meats like ham, turkey, and pastrami) (g/day) |
| **Red meat** | Consumption of red meat (includes beef, pork, lamb, and goat but excludes poultry, fish, eggs, and all processed meats) (g/day) |
| **Rice** | Rice availability per capita (g/day), a covariate |
| **Seafood omega-3 fatty acids** | Seafood omega-3 fatty acids (eicosapentaenoic acid and docosahexaenoic acid) in tablet or fish form (g/day) |
| **Second-hand smoke** | Average daily exposure to air particulate matter from second-hand smoke with an aerodynamic diameter smaller than $2.5 \mu\text{g}$, measured in $\mu\text{g}/\text{m}^3$, among non-smokers |
| **Smoking** | Prevalence of current use of any smoked tobacco product and prevalence of former use of any smoked tobacco product; among current smokers, cigarette equivalents smoked per smoker per day and cumulative pack-years of exposure; among former smokers, number of years since quitting |
| **Socio-demographic index** | SDI is a composite indicator of development status that was originally constructed for GBD 2015 and is derived from components that correlate strongly with health outcomes. |
|  | It is the geometric mean for indices of the total fertility rate among women younger than 25 years, mean education for those aged 15 years or older, and lag-distributed income per capita. The resulting metric ranges from 0 to 1, with higher values corresponding to higher levels of development. |
| **Sugar-sweetened beverages** | Consumption of any beverage with $\geq 50$ calories of sugar per one-cup serving, including carbonated beverages, sodas, energy drinks, fruit drinks but excluding 100% fruit and vegetable juices (g/day) |
| **Sweet potatoes** | Sweet potato availability per capita (g/day), a covariate |
| **Systolic blood pressure** | Systolic blood pressure (mm Hg) |
| **Total sugar** | Total sugar availability per capita (g/day), a covariate |
| **Vegetables** | Consumption of frozen, cooked, canned, or dried vegetables (including legumes but excluding salted or pickled, juices, nuts and seeds, and starchy vegetables such as potatoes or corn) (g/day) |
| **Vitamin A deficiency** | Proportion of children aged 0–5 years with serum retinol concentration $< 0.7 \mu\text{mol/L}$ |
| **Whole grains** | Consumption of whole grains (bran, germ, and endosperm in their natural proportions) from breakfast cereals, bread, rice, pasta, biscuits, muffins, tortillas, pancakes, and others (g/day) |

**Supplementary Table 2.** Omega-3 Fatty Acid g to fish g calculation¶.

| **Fish** | **DHA**<br>**g/3-ounce**<br>**fish** | **EPA**<br>**g/3-**<br>**ounce**<br>**fish** | **Omega-3 Fatty**<br>**Acids (DHA _**<br>**EPA) g/3-ounce**<br>**fish mean** | **Fish 3**<br>**ounces =**<br>**85.02 g** | **Fish (g) per**<br>**omega-3 Fatty**<br>**Acids**<br>**(g)=85.02 /**<br>**0.7264** |
| --- | --- | --- | --- | --- | --- |
| **Salmon Atlantic farmed** | 1.24 | 0.59 |  |  |  |
| **Salmon Atlantic wild** | 1.22 | 0.35 |  |  |  |
| **Herring Atlantic** | 0.94 | 0.77 |  |  |  |
| **Sardines canned in tomato sauce**<br>**drained** | 0.74 | 0.45 |  |  |  |
| **Mackerel Atlantic** | 0.59 | 0.43 |  |  |  |
| **Salmon pink canned drained** | 0.63 | 0.28 |  |  |  |
| **Trout rainbow wild** | 0.44 | 0.40 |  |  |  |
| **Oysters eastern wild** | 0.23 | 0.30 |  |  |  |
| **Sea bass** | 0.47 | 0.18 |  |  |  |
| **Shrimp** | 0.12 | 0.12 |  |  |  |
| **Lobster** | 0.07 | 0.10 |  |  |  |
| **Tuna light canned in water**<br>**drained** | 0.17 | 0.02 |  |  |  |
| **Tilapia** | 0.11 |  |  |  |  |
| **Scallops** | 0.09 | 0.06 |  |  |  |
| **Cod Pacific** | 0.1 | 0.04 |  |  |  |
| **Tuna yellowfin** | 0.09 | 0.01 |  |  |  |
| **Mean DHA and EPA Omega-3**<br>**Fatty Acids g/3 ounce fish** | 0.4531 | 0.2733 |  |  |  |
| **Calculations total Omega-3 FA g**<br>**to fish g** |  |  | 0.7264 | 85.02 | 117.043 |

¶ Data on omega-3 fatty acid content of varieties of fish came from the National Institutes of Health Office of Dietary Supplements (USA)

**Supplementary Table 3.** Calculations of KC/d from g/day of animal and plant foods¶.

| **Foods** | **Food sub-categories** | **Mean kcal/serving** | **Mean g/serving** | **Mean kcal/g** |
| --- | --- | --- | --- | --- |
| **Milk (2% fat)** |  | 122 | 244 | 0.5 |
| **Fish** |  | 218 | 170 | 1.28 |
| **Eggs** |  | 72 | 50 | 1.44 |
| **Poultry** |  | 187 | 85 | 2.91 |
| **Red meat** |  | 247 | 85 | 2.91 |
| **Processed meat** |  |  |  |  |
|  | **Salami** | 222 | 59 | 3.76 |
|  | **Pastrami** | 104 | 71 | 1.46 |
|  | **Ring baloney** | 86 | 28 | 3.07 |
|  | **Pepperoni** | 94 | 100 | 0.94 |
| **Average processed meat** |  | 126.5 | 64.5 | 1.96 |
| **Fruits** |  | 97 | 162 | 0.60 |
| **Vegetables** |  | 59 | 91 | 0.65 |
| **Legumes** |  | 249 | 179 | 1.39 |
| **Nuts** |  | 172 | 28 | 6.14 |
| **Seeds** |  |  |  |  |
|  | **Flax seeds** | 55 | 10 | 5.5 |
|  | **Chia seeds** | 58 | 12 | 4.83 |
|  | **Fennel seeds** | 34.5 | 10 | 3.45 |
|  | **Hemp seeds** | 55.3 | 10 | 5.53 |
| **Average of seeds** |  | 50.7 | 10.5 | 4.83 |
| **Average of nuts and seeds** |  | 111.4 | 19.25 | 5.78 |
| **Corn** |  | 99 | 103 | 0.96 |
| **Potatoes** |  | 161 | 173 | 0.93 |
| **Sweet potatoes** |  | 115 | 151 | 0.76 |
| **Rice** |  | 205 | 158 | 1.3 |
| **Whole grains** |  | 120 | 52 | 2.31 |

¶ Source: NutritionIX app<sup>12</sup>

## References

1. Teicholz N. The scientific report guiding the US dietary guidelines: is it scientific? BMJ. 2015; 351. http://www.bmj.com/bmj/351/bmj.h4962.full.pdf

2. Teicholz N. The scientific report guiding the US dietary guidelines: is it scientific? Rapid responses. BMJ. 2015; 351: h4962. https://www.bmj.com/content/351/bmj.h4962/rapid-responses

3. Global Burden of Disease Study 2017 (GBD 2017) Data Resources. Seattle, Washington: Institute of Health Metrics and Evaluation 2018; Accessed. http://ghdx.healthdata.org/gbd-2017

4. Global Health Data Exchange. Seattle, WA: Institute of Health Metrics and Evaluation 2017; Accessed January 10, 2020. http://ghdx.healthdata.org/

5. Naghavi M, Abajobir AA, Abbafati C, et al. Global, regional, and national age-sex specific mortality for 264 causes of death, 1980–2016: a systematic analysis for the Global Burden of Disease Study 2016. The Lancet. 2017; 390 (10100): 1151–1210. https://doi.org/10.1016/S0140-6736(17)32152-9

6. Flaxman AD, Lee YY, Vos T, et al. An Integrative Metaregression Framework for Descriptive Epidemiology. Seattle, WA: University of Washington Press, 2015.

7. GBD 2017 Risk Factor Collaborators. Global, regional, and national comparative risk assessment of 84 behavioural, environmental and occupational, and metabolic risks or clusters of risks for 195 countries and territories, 1990–2017: a systematic analysis for the Global Burden of Disease Study 2017: Table of risk factor definitions. The Lancet. 2018; 392 (10159): 1923–94. https://www.thelancet.com/action/showFullTableHTML?isHtml=true&tableId=tbl1&pii=S0140-6736%2818%2932225-6

8. PROTOCOL FOR THE GLOBAL BURDEN OF DISEASES, INJURIES, AND RISK FACTORS STUDY (GBD) Version 3.0; Issue 26. Seattle, Washington: Institute for Health Metrics and Evaluation Accessed August 2, 2019. http://www.healthdata.org/sites/default/files/files/Projects/GBD/GBD_Protocol.pdf

9. Food and Agriculture Organization of the United Nations. United Nations Accessed August 26, 2021. http://www.fao.org/faostat/en/#data

10. Call for Collaborators. Seattle, Washington: Institute for Health Metrics and Evaluation at the University of Washington 2019; Accessed April 25, 2019. http://www.healthdata.org/gbd/call-for-collaborators

11. Omega 3 Fatty Acids: Fact Sheet for Health Professionals. The Office of Dietary Supplements 2018; Accessed September 1, 2018. https://ods.od.nih.gov/factsheets/Omega3FattyAcids-HealthProfessional/

12. Nutritionix Track App. Syndigo LLC Accessed April 25, 2019. https://www.nutritionix.com/

13. Ritchie H, Roser M. Diet Compositions. Our World in Data. 2017. https://ourworldindata.org/diet-compositions

14. Willett W, Rockström J, Loken B, et al. Food in the Anthropocene: the EAT-Lancet Commission on healthy diets from sustainable food systems. The Lancet. 2019; 393 (10170): 447–492. https://doi.org/10.1016/S0140-6736(18)31788-4

15. Noncommunicable diseases: Mortality Geneva, Switzerland: World Health Organization 2021; Accessed August 25, 2021. https://www.who.int/data/gho/data/themes/topics/topic-details/GHO/ncd-mortality

16. Zagmutt FJ, Pouzou JG, Costard S. The EAT-Lancet Commission: a flawed approach? The Lancet. 2019; 394 (10204): 1140–1141. https://doi.org/10.1016/S0140-6736(19)31903-8

17. Torjesen I. WHO pulls support from initiative promoting global move to plant based foods. BMJ. 2019; 365: l1700. https://www.bmj.com/content/bmj/365/bmj.l1700.full.pdf

18. Willett W, Rockström J, Lang T, et al. Healthy Diets From Sustainable Food Systems, Food Planet Health: Target 1 Healthy Diets pg 10. Stockholm, Sweden: EAT-- Established by the Stordalen Foundation, Stockholm Resilience Centre and Wellcome Trust 2019; Accessed July 10, 2020. https://eatforum.org/content/uploads/2019/01/EAT-Lancet_Commission_Summary_Report.pdf

19. Beulens JWJ, Booth SL, van den Heuvel EGHM, Stoecklin E, Baka A, Vermeer C. The role of menaquinones (vitamin K2) in human health. British Journal of Nutrition. 2013; 110: 1357–1368. https://www.cambridge.org/core/journals/british-journal-of-nutrition/article/the-role-of-menaquinones-vitamin-k2-in-human-health/5B9F317B526629D8BA77B6435F1E5509

20. Zile MH. Vitamin A-not for your eyes only: requirement for heart formation begins early in embryogenesis. Nutrients. 2010; 2 (5): 532–50. https://www.ncbi.nlm.nih.gov/pmc/articles/PMC3257662/

21. Xing X, Tao F. [Advance of study on vitamin A deficiency and excess associatied with congenital heart disease]. Wei Sheng Yan Jiu. 2008; 37 (6): 754–6. https://pubmed.ncbi.nlm.nih.gov/19239019/

22. Saremi A, Arora R. Vitamin E and cardiovascular disease. Am J Ther. 2010; 17 (3): e56–65.

23. Wang L, Song Y, Manson JE, et al. Circulating 25-Hydroxy-Vitamin D and Risk of Cardiovascular Disease: A Meta-Analysis of Prospective Studies. Circulation: Cardiovascular Quality and Outcomes. 2012; 5 (6): 819–829. http://circoutcomes.ahajournals.org/content/5/6/819.abstract

24. Fu X, Harshman SG, Shen X, et al. Multiple Vitamin K Forms Exist in Dairy Foods. Current developments in nutrition. 2017; 1 (6): e000638–e000638. https://www.ncbi.nlm.nih.gov/pmc/articles/PMC5998353/

25. Walther B, Chollet M. Vitamin K2 - Vital for Health and Wellbeing. In: Gordeladze JO, ed. Menaquinones, Bacteria, and Foods: Vitamin K2 in the Die: IntechOpen, 2017.

26. Kamao M, Suhara Y, Tsugawa N, et al. Vitamin K content of foods and dietary vitamin K intake in Japanese young women. J Nutr Sci Vitaminol (Tokyo). 2007; 53 (6): 464–70. https://pubmed.ncbi.nlm.nih.gov/18202532/

27. Kaneki M HS, Hosoi T, Fujiwara S, Lyons A, Crean SJ, Ishida N, Nakagawa M, Takechi M, Sano Y, Mizuno Y, Hoshino S, Miyao M, Inoue S, Horiki K, Shiraki M, Ouchi Y, Orimo H. Japanese fermented soybean food as the major determinant of the large geographic difference in circulating levels of vitamin K2: possible implications for hip-fracture risk. Nutrition. 2001; 17 (4): 315–21. https://pubmed.ncbi.nlm.nih.gov/11369171/

28. Joseph M. Fermented Soy Products: A Guide To 12 Traditional Foods. Nutrition Advance 2020; Accessed April 11, 2021. https://www.nutritionadvance.com/fermented-soy-products/

29. Tamang JP, Cotter PD, Endo A, et al. Fermented foods in a global age: East meets West. COMPREHENSIVE REVIEWS IN FOOD SCIENCE AND FOOD SAFETY. 2020; 19 (1): 184–217. https://onlinelibrary.wiley.com/doi/full/10.1111/1541-4337.12520

30. Pu J, Romanelli R, Zhao B, et al. Dyslipidemia in special ethnic populations. Cardiology clinics. 2015; 33 (2): 325–333. https://www.ncbi.nlm.nih.gov/pmc/articles/PMC4421090/

31. Frank ATH, Zhao B, Jose PO, Azar KMJ, Fortmann SP, Palaniappan LP. Racial/ethnic differences in dyslipidemia patterns. Circulation. 2014; 129 (5): 570–579. https://www.ncbi.nlm.nih.gov/pmc/articles/PMC4212818/

32. Keys A, Menotti A, Karvonen MJ, et al. The diet and 15-year death rate in the seven countries study. Am J Epidemiol. 1986; 124 (6): 903–15. https://academic.oup.com/aje/article-abstract/124/6/903/174332?redirectedFrom=fulltext

33. Mahmood SS, Levy D, Vasan RS, Wang TJ. The Framingham Heart Study and the epidemiology of cardiovascular disease: a historical perspective. Lancet (London, England). 2014; 383 (9921): 999–1008. https://www.ncbi.nlm.nih.gov/pmc/articles/PMC4159698/

34. Dehghan M, Mente A, Zhang X, et al. Associations of fats and carbohydrate intake with cardiovascular disease and mortality in 18 countries from five continents (PURE): a prospective cohort study. The Lancet. 2017; 390 (10107): 2050–2062. https://doi.org/10.1016/S0140-6736(17)32252-3

35. Countries Who Eat The Most Cheese. World Atlas 2021; Accessed March 28, 2021. https://www.worldatlas.com/articles/countries-who-consume-the-most-cheese.html#:~:text=The%20top%20cheese%20consumer%20is,kilograms%20of%20cheese%20per%20capita.

36. Cheese Consumption Per Capita in New Zealand. Czech Republic: Helgi Library, Source: Faostat Accessed April 9, 2021. https://www.helgilibrary.com/indicators/cheese-consumption-per-capita/new-zealand/

37. Khalil Z, Alam B, Akbari AR, Sharma H. The Medical Benefits of Vitamin K(2) on Calcium-Related Disorders. Nutrients. 2021; 13 (2): 691. https://www.ncbi.nlm.nih.gov/pmc/articles/PMC7926526/

38. POTATO PROCESSING AND USES. Lima, Peru: International Potato Center Accessed May 28, 2020. https://cipotato.org/potato/potato-processing-uses/

39. Juul F, Vaidean G, Lin Y, Deierlein Andrea L, Parekh N. Ultra-Processed Foods and Incident Cardiovascular Disease in the Framingham Offspring Study. Journal of the American College of Cardiology. 2021; 77 (12): 1520–1531. https://doi.org/10.1016/j.jacc.2021.01.047

40. Srour B, Fezeu LK, Kesse-Guyot E, et al. Ultra-processed food intake and risk of cardiovascular disease: prospective cohort study (NutriNet-Santé). BMJ. 2019; 365: l1451. https://www.bmj.com/content/bmj/365/bmj.l1451.full.pdf

41. Monteiro CA, Moubarac JC, Cannon G, Ng SW, Popkin B. Ultra-processed products are becoming dominant in the global food system. Obes Rev. 2013; 14 Suppl 2: 21–8. https://pubmed.ncbi.nlm.nih.gov/24102801/

42. Mozaffarian D, Fahimi S, Singh GM, et al. Global Sodium Consumption and Death from Cardiovascular Causes. New England Journal of Medicine. 2014; 371 (7): 624–634. https://www.nejm.org/doi/full/10.1056/NEJMoa1304127

43. O’Donnell M, Mente A, Yusuf S. Sodium Intake and Cardiovascular Health. Circulation Research. 2015; 116 (6): 1046–1057. https://www.ahajournals.org/doi/abs/10.1161/CIRCRESAHA.116.303771

44. Roseboom T, van der Meulen JHP, Osmond C, et al. Coronary heart disease after prenatal exposure to the Dutch famine, 1944-45. Heart. 2000; 84 (6): 595–598. https://www.heart.bmj.com/content/84/6/595

45. Dieterich CM, Felice JP, O’Sullivan E, Rasmussen KM. Breastfeeding and health outcomes for the mother-infant dyad. Pediatric clinics of North America. 2013; 60 (1): 31–48. https://www.ncbi.nlm.nih.gov/pmc/articles/PMC3508512/

46. Ambient (outdoor) air pollution. Geneva, Switzerland: World Health Organization 2018; Accessed March 24, 2021. https://www.who.int/news-room/fact-sheets/detail/ambient-(outdoor)-air-quality-and-health

47. Tobacco responsible for 20% of deaths from coronary heart disease Geneva, Switzerland: World Health Organization 2020; Accessed March 2, 2021. https://www.who.int/news/item/22-09-2020-tobacco-responsible-for-20-of-deaths-from-coronary-heart-disease

48. Vaziri ND. Mechanisms of lead-induced hypertension and cardiovascular disease. American Journal of Physiology-Heart and Circulatory Physiology. 2008; 295 (2): H454–H465. https://journals.physiology.org/doi/abs/10.1152/ajpheart.00158.2008

49. Lanphear BP, Rauch S, Auinger P, Allen RW, Hornung RW. Low-level lead exposure and mortality in US adults: a population-based cohort study. The Lancet Public Health. 2018; 3 (4): e177–e184. https://doi.org/10.1016/S2468-2667(18)30025-2

50. Bikbov B, Purcell CA, Levey AS, et al. Global, regional, and national burden of chronic kidney disease, 1990-2017: a systematic analysis for the Global Burden of Disease Study 2017. The Lancet. 2020; 395 (10225): 709–733. https://doi.org/10.1016/S0140-6736(20)30045-3

51. Global, regional, and national age-sex-specific mortality for 282 causes of death in 195 countries and territories, 1980-2017: a systematic analysis for the Global Burden of Disease Study 2017. Lancet. 2018; 392 (10159): 1736–1788. https://www.thelancet.com/journals/lancet/article/PIIS0140-6736(18)32203-7/fulltext

52. Afshin A, Sur PJ, Fay KA, et al. Health effects of dietary risks in 195 countries, 1990-2017: a systematic analysis for the Global Burden of Disease Study 2017. The Lancet. 2019; 393 (10184): 1958–1972. https://www.thelancet.com/journals/lancet/article/PIIS0140-6736(19)30041-8/fulltext

53. Stevens GA, Alkema L, Black PRE, et al. Guidelines for Accurate and Transparent Health Estimates Reporting: the GATHER statement. The Lancet. 2016; 388 (10062): e19–e23. https://doi.org/10.1016/S0140-6736(16)30388-9

